# A qualitative study exploring the experience of the Medical Intern Programme: Supportive transition for international doctors working in the NHS

**DOI:** 10.1101/2021.06.14.21258746

**Authors:** J. F. Lavallée, S. Conen, D. R. Corfield, J. Howells, M. Pugh, J. Hart

**Affiliations:** Division of Medical Education, University of Manchester, UK; Lancashire Teaching Hospitals, UK

**Keywords:** international workforce, health service, recruitment and retention

## Abstract

**Background:** The UK is experiencing a shortage of doctors. Consequently, the Medical Intern Programme, a unique two year programme consisting of an observership, four clinical rotations at the level of a foundation doctor within an NHS Trust and a postgraduate diploma from a university in the north of England, was set up to enhance the NHS workforce by facilitating the entry of international medical graduates into UK medicine via a supported transitional programme. We aimed to explore the experiences of the doctors enrolled on the Medical Intern Programme.

**Methods:** Semi-structured interviews were conducted via the telephone with seven doctors enrolled on the programme. The interview guide was informed by the research questions and data were analysed using a thematic analysis.

**Results:** We identified four themes that were important in the experiences of the doctors: *preparing to work in the UK, feeling supported, weighing up the pros and cons of the programme and comparisons between countries*.

**Conclusions:** The Medical Intern Programme successfully facilitated international doctors’ transition to the UK and working in the NHS. Support needs to be provided at the organisational, team and individual level including a period of observing and shadowing to enable the doctors to understand the systems and multidisciplinary team working in the NHS.

## Background

Healthcare workers are individuals who engage in actions to enhance health (World Health Organisation, 2016). Globally, healthcare is an important aspect of society and one of the largest employers; it is estimated to be worth over $9 trillion worldwide (Britnell, 2019) and contributes to national economies. In the UK, approximately 2.8 million people are employed within health and social care (Horton, et al., 2016). However, the UK faces a longstanding challenge of educating, training and retaining healthcare workers; and the percentage of doctors who move from foundation training directly into speciality training has reduced from 71% in 2011 to 34.9% in 2019 (The Foundation Programme, 2019), although some may later return to training. The King’s Fund have suggested that staff shortages across the NHS in England alone could reach 350,000 by 2030 (The King’s Fund, 2018). Consequently, there has been a reliance upon the international recruitment of healthcare workers (World Health Organisation, 2019; Moon & Shin, 2018).

Over the past decade, there have been a number of challenges that prompted global efforts to respond to health emergencies and prevent future crises (e.g., COVID-19, Zika virus, Ebola). Additionally, the workforce is being viewed as global with many countries now being seen as the source and destination of healthcare workers (World Health Organisation, 2019). The World Health Organisation reported a 60% rise in the number of migrant doctors and nurses over the last decade (World Health Organisation, 2019). There are a number of common ‘push’ and ‘pull’ factors that influence migration for healthcare workers. ‘Push’ factors are those that are internal to the country and encourage the person to move away either to a different part of the country or from their home country; and ‘pull’ factors are external to the person’s home country and are attributes that attract individuals. Common push and pull factors include financial factors, professional development, working conditions and general socio-political factors (Cleland et al., 2016; Nair & Webster, 2013; Takemura et al., 2016), as well as mobility factors such as visa procedures and active recruitment strategies (Kovacs et al., 2014; Sheikh et al., 2012).

There are a number of challenges with migration on both an individual and global level. Individuals often face difficulties in adjusting and transitioning to the culture of their host country and healthcare system (Bond et al., 2020). Global public health has recognised the difficulties posed by healthcare workers moving from low- and middle-income countries to high-income countries (Oladeji & Gureje, 2016). Migration has had a significant impact on the quality of healthcare systems in source countries and is a threat to achieving health-related sustainable development goals (World Health Organisation, 2016). Moreover, areas with greater disease burden have lower numbers of healthcare workers, exacerbating the impact of the healthcare worker deficit.

Despite the difficulties of migration for healthcare workers, there are a number of benefits for the receiving country and potential global benefits too. Migration enables the workforce to effectively navigate different cultures and healthcare systems, whilst building global attributes and skillsets; ultimately developing a global mind set in line with sustainable development goals. Thus, the benefits of recruiting international doctors are broader than only reducing the workforce deficit. An additional benefit includes having a workforce that reflects the diversity of the UK population.

In 2020, an NHS Trust in the north of England partnered with a University and developed a two year Medical Intern Programme (MIP) to recruit international doctors from one low- and middle-income country in the first instance. Eligibility criteria for the MIP were a good medical degree from a recognised University/medical course similar in structure to a UK undergraduate medical degree course, plus General Medical Council (GMC) registration/Professional Linguistic Assessment Board (PLAB). The aim of the MIP was to reduce some of the workforce deficits, attract the best quality international doctors to the UK and to retain these doctors through professional development in preparation for future training posts. The training needs of international doctors have informed the development of the MIP. The MIP consists of supported recruitment, appointment/visa processes, support with settling in (e.g., advice regarding accommodation, finances), peer group support, a bespoke induction, career development discussions, an observership, four clinical rotations as a foundation doctor within an NHS Trust, a part-time two year postgraduate diploma and eligibility to apply for speciality training in the UK on completion of the course. The diploma is led by the university, with part of the teaching delivered at the NHS trust. Doctors have some protected time for study. Modules include a focus on the NHS, patient-centredness, governance, professionalism and specific clinical practice development. Optional modules include medical education, health services research, policy and management and public health. Often doctors recruited from low- and middle-income countries are unable to access postgraduate courses whilst working with a Tier 2 visa (Trewby, 2017). Therefore, this is a unique training opportunity for international doctors to work in the NHS and experience a number of different rotations whilst studying for a post-graduate university qualification. We aimed to explore the experiences of the doctors on the MIP to understand their perspectives on working in the UK and the programme.

## Methods

### Study design

We conducted a qualitative study using semi-structured interviews.

### Participants

At study onset there were nine doctors enrolled in the programme. We used purposive sampling to recruit doctors employed though the MIP at an NHS hospital in the north of England and registered on the Diploma at the partner university. An invitation email explaining the purpose of the study was sent to potential participants, along with a Participant Information Sheet. Those who were interested in taking part were invited to contact the researcher who was not involved in the delivery of the MIP (JL). If individuals agreed to take part, verbal consent was gained. Sample size was informed by the number of doctors enrolled onto the MIP.

### Data collection

Due to the COVID-19 global pandemic and restrictions in place, one researcher conducted the semi-structured interviews via the telephone (JL). The interviews were informed by the research aim and included questions such as *“why did you choose to apply for the programme?”, “how has your experience of moving country been?”* and *“how did you find the observership?”* We did not collect demographic data as the cohort was small and we wanted to ensure anonymity for all participants. Interviews were audio-recorded, transcribed verbatim and proof-read. Names were removed from all of the transcripts to ensure anonymity and the participants will be referred to as ‘doctor’ followed by a non-identifiable number.

### Data analysis

We managed the data in NVivo 12 and analysed the data inductively using Thematic Analysis (Braun & Clarke, 2006). All transcripts were analysed by one author (JL) and SC and JH independently reviewed a proportion of the transcripts. Any discrepancies were discussed and resolved.

### Ethics

This study was approved by the standing proportionate University Research Ethics Committee. Participants gave their verbal informed consent prior to study participation.

### Findings

A total of seven doctors took part in semi-structured telephone interviews lasting between 29 and 41 minutes (average of 33 minutes). The remaining two doctors did not respond to the researcher. During the inductive analysis, four themes emerged from the data: *(i) preparing to work in the UK, (ii) feeling supported, (iii) weighing up the pros and cons of the programme, and (iv) comparisons between countries*.

### Preparing to work in the UK

All participants were already planning to work in the UK and had gained their GMC registration and completed their PLAB test. Many of the participants were actively applying for jobs in the UK when they heard about the MIP. One participant explained that they had been applying for jobs in the UK for five years. The number of job applications submitted exceeded over 100 for most of the participants, yet they explained that they had not been shortlisted for interviews.

> *“after I finished my GMC registration I was looking for a job in the UK. So I applied for 100, maybe 150 jobs, by the time that the announcement for the programme came out*.*” (Doctor, 2)*

Two of the MIP stakeholders visited the doctors’ home country to deliver a seminar about the programme but doctors from the current cohort did not attend this; some did not know about the seminar and others thought it was a postgraduate diploma only. The doctors heard about the details of the programme through friends, University tutors and Facebook. The doctors explained that those who were eligible to apply for the programme did apply.

### Feeling supported

#### Application

The doctors explained that the application process was straightforward and similar to other NHS job applications, making the process of applying relatively easy. One participant spoke about the support they received in completing their paperwork, which was new to them, and how quickly Human Resources responded to emails, helping them to feel supported throughout the process. The participants explained that having enough time during the interview was important as sometimes they needed to ask questions for clarity or took a little longer as the interview was conducted in English which was not their native language.

> *“But then he was really, really lovely. He was very; he was really detailed in his questions. He gave me all the time I needed to ask my questions at the end*.*” (Doctor, 7)*

However, all of the doctors explained that they would have liked more information about the postgraduate diploma, rotations and University fees whilst applying for the programme. This would have enabled them to feel more secure in their decisions, but they understood that this information was not available at the time of interview as this was a new course.

#### Peers and the community

The doctors thought that they settled into life in the UK a lot easier because they came with other international doctors from their home country. Supporting each other and sharing similar experiences was invaluable to the doctors. Some of them were close friends back in their home country. Moreover, being placed in a northern city where there was a diverse population helped some of the doctors to settle in as they felt able to be part of the community, especially in regards to their religious beliefs.

> *“It’s not difficult for me, because of my friends here, so we can, I mean we have familiar people with us. But if I was alone here maybe it would be more difficult. But generally speaking living in the UK is easy. Especially first time because of our religious beliefs and there are a lot of diversity here in [UK city], so it made it easier for us to go fast [settle] in the community*.*” (Doctor, 5)*

#### Medical Intern Programme

All of the doctors spoke about how well supported they felt from the stakeholders involved in the programme. Most of the doctors first moved to the UK in August 2020 during the COVID-19 pandemic and this provided additional challenges as the doctors were required to isolate for 14 days on arrival and some shops and local amenities were not open as usual. The local stakeholders provided shopping for the doctors for the first 14 days and then took them on a tour of the local area. In addition, the doctors explained that the “system” here was confusing and different to their home country so receiving help with daily administrative tasks such as setting up bank accounts was helpful. The doctors stated that the team went above and beyond to help them settle in and this made a big difference to them. They did not expect to be treated in this way, but it made them feel very welcome and supported.

> *“I expected people to be professional. I expected them to be pragmatic, but I didn’t expect them to treat someone that they don’t really know that well, to treat them that well. So it was something that really touched my heart. It was really good*.*” (Doctor, 7)*

The doctors explained that they met weekly with the MIP lead and this was very helpful. The doctors felt supported as they could talk to the stakeholder about some of the challenges they were experiencing. For example, one doctor explained that they had experienced some difficulties with a colleague on the ward and the stakeholder was able to help with this situation. Being able to discuss a variety of issues with the stakeholder was described as “a relief” by one doctor. In addition, the doctors explained that if they were finding any aspects of the diploma difficult, they could speak with the tutors and knew that they would receive the support they needed.

The doctors explained that the learning environment for the diploma was a supportive one where they believe their opinions matter and are provided with the space to express their thoughts. One doctor explained that it does not feel like teaching, rather *“it is like talking with a wise friend”* enabling a deeper reflection about the learning.

#### Observership

Before the doctors could begin working in the NHS Trust, they completed the mandatory Trust induction online. The doctors explained that they found only some aspects of the Trust induction helpful such as equality and diversity, but wanted to know more about their role as a junior doctor and how “the system” works. Instead, the doctors learned by asking and shadowing colleagues during the observership.

The observership provided the doctors with one month of shadowing other staff members prior to working as a doctor, allowing them to observe practices and learn about It systems and ward processes. The observership provided the doctors with the time and opportunity to introduce themselves to their colleagues and learn about the different roles within the multidisciplinary team. The doctors explained that in their home country the healthcare team was made up of doctors and nurses. As a consequence, the doctors were unfamiliar with the roles of allied health professionals such as physiotherapists, occupational therapists and speech and language therapists.

The observership helped the doctors to feel more supported during the transition of moving from one healthcare system to a new one. Initially, the doctors observed colleagues. Some of the doctors explained that the observership was one of the reasons they applied for the MIP. They had heard from peers working in the UK that it was difficult to adapt to working in the NHS. Consequently, the doctors thought that this supported period would be beneficial for them. The observership provided the doctors with the time and space to ask lots of questions and they were gradually given more responsibility which helped to increase their confidence, especially with the computer systems.

> *“The shadowing period, yes, so it was very important for us. And it was very useful because it’s the first job in UK. We are ok, we are doctors, so we have worked with patient before but here in the UK it’s not only about being a doctor, it’s about fitting in the system and doing things in the right way*.*” (Doctor, 3)*

Initially, the observership was planned to last for two months, but this time was reduced to one month as the doctors adapted more quickly than expected to working in the NHS.

The quality of the observership appeared to be influenced by the availability of staff members. One doctor spoke about their experience of shadowing a locum doctor for the whole month, and this was problematic as they were not able to observe a range of people and this particular doctor worked very quickly.

> *“In my case it wasn’t really helpful because I had to shadow a locum who was more senior than I was, and he was the only doctor, so it was difficult for me to cope with him. He was really fast and I didn’t know anything about the system or the methods of nursing and so a lot of the things were different for me*.*” (Doctor, 5)*

Nevertheless, the doctors explained that both permanent and locum staff members provided them with the opportunity to learn about the NHS and the varied roles within it.

Some of the doctors were uncertain about whether the team on the ward was expecting them and thought their colleagues were not always aware that they were international doctors and new to working in the NHS. The doctors found it difficult to know who to shadow and what they needed to gain from the experience. Having clear learning objectives for the observership was suggested as a way to improve this. Finally, two doctors spoke about their concerns of how others in the team perceived their levels of competence due to needing to shadow for the month and that it was “disappointing” to not be helping.

#### Working in the NHS

The doctors explained that they believed the NHS was a supportive environment to work in where staff can speak with each other about the different treatment options, providing *“the opportunity to learn without compromising patient safety”*.

> *“Sometimes, people come and say “good job”, and you know this is really cool*.*” (Doctor, 6)*

The doctors were pleasantly surprised by the support for NHS staff when they arrived in the UK as they saw signs thanking the NHS and were given discounts when buying items. They believe that

> *“everyone are trying to help each other, they are trying to improve systems, trying to improve the quality of care to the patient*.*” (Doctor, 2)*.

The doctors were surprised by how much paperwork there was to complete when working in the NHS and it took a while for them to get used to this. They recognised the importance of completing the paperwork, in following the guidelines for procedures and always felt supported in following these procedures.

> *“everyone is trying to stick to what is written. So they have to do a procedure in a specific way, they will do it in the way it’s written. There is always people who supervise what you are doing, so there is no, there is slight space for mistakes, from the human effect” (Doctor, 2)*

All of the doctors discussed how much they enjoyed working in the NHS. They really appreciated having a sufficient amount of resource to use and how these were applied equally across patients depending on their needs. They explained that the system does not discriminate against who gets treatment and who does not, as *“every patient has got the right for the highest care”*. They believed that the NHS as an organisation really cares for people and offers the best care available.

> *“There is no limitations for what you can do for the patient, that’s very important thing so every patient who is healthy, not healthy, every patient has got the right for the highest care, the highest delivery of escalation. Whatever his or her disease is, yeah that’s really important and the doctors are always supported” (Doctor, 3)*

### Weighing up the pros and cons of the programme

#### Undertaking a postgraduate diploma

The reputation of the University hosting the programme was an attractive aspect for the doctors. The UK University already had well-established links with the University they trained at in their home country and this was helpful for some of the doctors, but others thought it would be beneficial if the programme was open for doctors from other Universities in their home country.

##### Financial implications

The doctors spoke about the financial implications of undertaking the postgraduate diploma. The doctors were not aware of how much the course fees would be prior to starting the programme. Once the course fees had been communicated, the doctors engaged in conversations about how to pay and it was agreed that the fees would be deducted from their salary each month. They explained that it was a burden for them to find the money to pay the course fees each month and at the beginning it was challenging to plan for because they did not know what their take home salary would be.

The course fees were described as a “financial investment” by one doctor as they believed the postgraduate diploma would be helpful for their future. Another doctor explained that they did not come to the UK to save money, but wanted to know that they could afford rent and living costs. The doctors’ uncertainty regarding their finances was made more difficult as the doctors had to wait some time before receiving their initial salary payment. The financial implications was an important factor that influenced whether or not the doctors would recommend the programme to others.

> *“one of my colleagues, if his main priority is saving money, he wouldn’t come here, and he would go with any Trust or service job. Like he would do the same amount of work, and the same hours and get I think a fair amount of money more. So for this I wouldn’t recommend coming here*.*” (Doctor, 6)*

##### Personal and professional development

All of the doctors stated that the postgraduate diploma was one of the reasons they applied for the MIP. The doctors explained that the diploma provided them with the opportunity to develop their knowledge of the NHS and critical appraisal skills. These skills were helpful in their work for a number of reasons including facilitating their knowledge of a different healthcare system and providing them with another focus that was separate but related to work. All of the doctors believed the postgraduate diploma added value to the MIP as they thought this would help them in their future roles and applications for speciality training (a Postgraduate Diploma may enhance the chances of a successful application for a Higher Specialist Training Post). One doctor described it as *“an offer you can’t refuse”*.

> *“I applied because my aim was to work in the NHS at first and thought it would be a good opportunity, not just the work but to have a diploma added to this and I knew this may help me in the future*.*” (Doctor, 2)*

Whilst the doctors saw the many benefits of completing the diploma, they also talked about the challenges of studying whilst working and explained that they were finding it difficult to meet deadlines and reach a work-life balance due to their shift patterns.

> *“it’s not easy to start a new job in a new country and try to adapt as much as you can in the job, and the same time you are trying to adapt like this postgraduate study because you don’t know about it as well. So you are trying to run in two ways, parallel together, so it’s not easy to do it but you feel the benefit of it*.*” (Doctor, 3)*

Moreover, the doctors discussed the disappointment they felt when they received the grade for their first assessment as their marks were much lower than they had expected and had received previously in their home country. The need to have appropriate expectations was important and there was an element of adjustment required by the doctors.

#### Length of Medical Intern Programme

The length of the programme was described as *“double edged”*. The doctors stated that it is quite a commitment and thought that others may not apply for the programme if their aim was to enter the training routes as quickly as possible. However, they explained that having two years of employment, rotations and experience within the NHS was beneficial as they were able to gain a lot of varied experiences; and one doctor felt that this was a safe option.

> *“I know this train will lead me to this destination, I think it might be a little bit slow, but if I choose a faster track I’m not sure what could happen” (Doctor, 6)*

The doctors explained that the rotations were an important aspect of the MIP and one of the reasons they applied. They explained that over the two year period they will work across four different specialities, providing them with lots of valuable experiences across medicine and surgery similar to the Foundation Programme that other NHS jobs would not provide. *“usually when you apply for some jobs you don’t get vocations, you just do the job for about one year or six months so that’s not a great exposure to the medical practice in the UK. While here I have 2 years of 4 different rotations so this is going to give me a lot more exposure than usual service jobs*.*” (Doctor, 5)*

However, the doctors explained that they would have liked more information about the rotations and the different jobs they would be working in prior to starting the programme. Initially the doctors ranked their preferred rotations, but found out which rotations they would be completing for their first year once they arrived in the UK. They also stated that they still did not know the rotations for their second year and this was something they thought could be improved for future cohorts. They also thought it would be helpful to understand how each doctor was assigned to their rotation.

One doctor explained that it was important for them to experience different specialities in other countries to enable them to make an informed decision about the area of medicine they would like to specialise in. All of the doctors agreed that the role of a doctor can change depending on the health service and country they are working in and two years was a good amount of exposure to the NHS.

> *“medicine is the same but the job is not the same at all” (Doctor, 4)*

#### Comparisons between countries

All of the doctors talked about the differences between England and their home country. There were two main comparisons made which the doctors explained had impacted on how quickly they settled in. Firstly, the doctors were surprised by the culture in England, in particular the timings of social activities.

> *“Usually we start our days in the evening. The evening is very (laughs) people go out and have a life. In the UK after 5, everything just shut down and you have to stay at home, so the life style of the people is completely different from ours and it gets time to get used to this and also getting the rests, is very difficult*.*” (Doctor, 2)*

Some of the doctors were uncertain about whether this was due to COVID and the restrictions in place, but they believed this to be a big difference and something that they would need to adapt to. The doctors explained that having a social life was very important to them and it helped them to feel more “energised”. They had expected to have a social life whilst living in England and they felt disappointed by this.

Secondly, the doctors talked about the weather in England and how much they missed the sunshine. This was something that they were having to adapt to, but were finding it difficult.

> *“It’s not the greatest weather in the world and coming from a country that is always sunny, I really, really miss the sun. It’s one of these things that I never expected to have to take vitamin D tablets every day because I don’t get to see the sun. Because I go to work before the sunrise and come home after the sunset. So I almost never see the sun” (Doctor, 7)*

One doctor spoke about the challenges of speaking another language as they thought their English was conversational. They explained that they did not find this problematic in the workplace as they believed medical terminology to be the same in all countries, but it was more problematic in their personal life when trying to meet new people. Another doctor explained that it was helpful to speak different languages as during their observership they took a history from a patient who only spoke Arabic and presented the information to a senior.

## Discussion

The MIP is a two year postgraduate programme in the UK offered to international doctors consisting of four rotations in the NHS and a postgraduate diploma. This study explored the doctors’ perspectives of the MIP and found a number of benefits, including providing professional development opportunities and supporting the transition of moving country and healthcare systems. The areas for improvement included understanding the cost of the postgraduate diploma and the amount of information available about the MIP.

More than a third of registered doctors in the UK have trained elsewhere (Webb et al., 2014). International doctors dedicate a lot of time to upskilling and completing the requirements to work abroad in countries such as the UK, Germany and the USA. Whilst the move to another country and healthcare system can be desirable, the transition is challenging and supporting doctors to make this transition is beneficial. Many international healthcare workers are often not provided with the opportunity to adapt their skills and knowledge in line with their host country (Ohr et al., 2014). It is well documented within the literature that many of the international workforce have difficulties with practical issues, as well as the structures and culture of the host country (Kehoe et al., 2016; Slowther et al., 2012), which may increase their risk of being referred for fitness to practice (Tiffin et al., 2014). Within the current study, the support provided to the international doctors was viewed as a unique opportunity. There was a team-based approach linked with strong leadership that facilitated the doctors’ transition to working in the UK and within the NHS. However, some doctors explained that they thought their colleagues were not always aware that they were international doctors and new to working in the NHS. Therefore, educating and supporting existing staff may also be important for the overall success of the transition for international healthcare workers.

Successful recruitment, transition and retention of international healthcare workers has been found to be influenced by organisational support (Ohr et al., 2014). Organisational support may be demonstrated in a number of ways. The doctors in the current study explained that being provided additional time in their interview was really helpful and having their needs recognised was an important factor in helping them to feel supported. Depending on the stage of their career, international doctors may need different interventions to support their needs. We have demonstrated that the observership and introduction to the NHS were really important aspects of the MIP and one that the doctors found helpful and attractive, even if they had already worked in the NHS. Induction programmes are implemented across NHS Trusts, but often there is insufficient consideration provided to the content of what is needed for international doctors and how long for. In addition, it would be impractical to try to adapt generic inductions to include specific needs of groups.

A realist review of 88 studies found that interventions were more likely to be successful if they targeted individual needs within a supportive learning environment where support was provided by both peers and supervisors (Kehoe et al., 2016). Kehoe et al. (2016) provide a number of recommendations for implementing interventions to support international doctors including having cultural awareness within a programme, individual needs assessment and the provision of information about work, culture and general issues at the earliest opportunity. Moreover, successful transitions are facilitated by social networks that provide support and exchange information (Schumann et al., 2019). Such social networks enable members to develop and maintain their own identity, as well as share the beliefs and practices important and relevant to their culture. Whilst the programme content and delivery are viewed as important, having an organisational culture that is welcoming and creates social support can have a big impact on how well international doctors transition into their host country (Kehoe et al., 2016). Interestingly, the participants in the current study wished to come to the UK and embark upon the MIP despite not knowing the details of the programme; suggesting that they perceived a number of attractive components within the MIP including the early organisational support during recruitment, the observership and the opportunity to complete a postgraduate diploma alongside rotations.

Recruiting and retaining international workforce is important for the NHS, but we need to recognise the potential impact that this has on low- and middle-income countries. The World Health Organisation has warned that there could be a global deficit of 750,000 doctors by 2030. Consequently, there is a need to focus on collaboration rather than competition (THET, 2019). However, the Medical Training Initiative “train and return” included within the NHS long-term plan requires further development as many international doctors plan to leave their home country and do not necessarily plan to return.

### Limitations

There are some limitations in this study. Firstly, the total number of doctors enrolled on the MIP was nine and all of the doctors were from the same country. Consequently, our sample size was small, but our sample size and findings were similar to other studies exploring interventions to recruit and retain international workforce (Al-Mohawes et al., 2021; Subedi et al., 2020). Secondly, we were not able to collect demographic information as this was a small sample and we prioritised anonymity. In future, it will be helpful to gather demographic information to learn more about the cohort and contextualise the findings further. Finally, we collected data from the doctors only. We plan to collect data from the doctors and stakeholders at different points across the next two years and this will enable us to take a holistic view of the MIP. It would also be beneficial to gather data from the Trust and staff who work alongside the international doctors to explore what support they may need.

## Conclusion

International doctors are keen to work in the UK and develop professionally. This study highlights a number of important factors when recruiting international doctors to the NHS. Firstly, the importance of delivering a programme where doctors’ professional developmental needs are recognised and supported. Secondly, the importance of encompassing additional support from the organisation and peers within programmes for international doctors. There are a number of additional needs that must be considered when recruiting and employing international doctors, and the MIP is one of the first programmes to address factors such as peer support, the social situations of the doctors, cultural differences and professional development.

## Data Availability

Data have been archived in line with ethics requirements.

## List of abbreviations

MIP: Medical intern programme
UK: United Kingdom
NHS: National Health Service
PLAB: Professional Linguistic Assessment Board
GMC: General Medical Council

## Declarations

### Ethics approval and consent to participate

This study was approved by the standing proportionate University of Manchester Research Ethics Committee.

### Consent for publication

All participants provided verbal consent in line with the ethics approval and using the University of Manchester’s consent form.

### Availability of data and materials

The datasets generated and/or analysed during the current study are not publicly available due to maintaining individual privacy but are available from the corresponding author on reasonable request.

### Competing interests

JL declares no competing interests

SC is a member of the University diploma team

DC is a member of the University diploma team and the MIP steering group

JH is a member of the MIP steering group

MP is MIP programme lead

JH is a member of the University diploma team and the MIP steering group

### Funding

No funding was received for this study.

## Authors’ contributions

**JL** designed the study, collected and analysed the data, drafted the manuscript and approved the submitted version. **SC** designed the study, analysed the data, contributed to the manuscript and approved the submitted version. **DC** contributed to the manuscript and approved the submitted version. **JH** contributed to the manuscript and approved the submitted version. **MP** contributed to the manuscript and approved the submitted version. **JH** designed the study, analysed the data, contributed to the manuscript and approved the submitted version.

## Acknowledgements

The authors would like to thank all of the participants for their contributions to this study.

## Notes

### Author Declarations

This study was approved by the standing proportionate University Research Ethics Committee at The University of Manchester. Participants gave their verbal informed consent prior to study participation.

## References

Al-Mohawes, H., Amante, M., Hannon, B., Zimmermann, C., Kaya, E., & Al-Awamer, A. (2021). Online Bridging Program for new international palliative medicine fellows: development and evaluation. BMJ Supportive and Palliative Care. https://doi.org/10.1136/bmjspcare-2020-002797

Bond, S., Merriman, C., & Walthall, H. (2020). The experiences of international nurses and midwives transitioning to work in the UK: A qualitative synthesis of the literature from 2010 to 2019. In International Journal of Nursing Studies. https://doi.org/10.1016/j.ijnurstu.2020.103693

Braun, V., & Clarke, V. (2006). Braun, V., Clarke, V. Using thematic analysis in psychology., 3:2 (2006), 77–101. Qualitative Research in Psychology.

Britnell, M. (2019). Human: Solving the global workforce crisis in healthcare. In Human: Solving the global workforce crisis in healthcare. https://doi.org/10.1093/oso/9780198836520.001.0001

Cleland, J., Johnston, P., Watson, V., Krucien, N., & Skåtun, D. (2016). What do UK doctors in training value in a post? A discrete choice experiment. Medical Education. https://doi.org/10.1111/medu.12896

Horton, R., Araujo, E., Bhorat, H., Bruysten, S., Jacinto, C., McPake, B., Reddy, K., Reinikka, R., Schmidt, J., Song, L., Tangcharoensathien, V., Trent-Adams, S., Weakliam, D. and Yamin, A. (2016). Final report of the expert group to the High-Level Commission on Health Employment and Economic Growth. Available at: http://www.world-psi.org/en/final-report-expert-group-high-level-commission-health-employment-and-economic-growth

Kehoe, A., McLachlan, J., Metcalf, J., Forrest, S., Carter, M., & Illing, J. (2016). Supporting international medical graduates’ transition to their host-country: realist synthesis. Medical Education. https://doi.org/10.1111/medu.13071

Kovacs, E., Schmidt, A. E., Szocska, G., Busse, R., McKee, M., & Legido-Quigley, H. (2014). Licensing procedures and registration of medical doctors in the European Union. Clinical Medicine, Journal of the Royal College of Physicians of London. https://doi.org/10.7861/clinmedicine.14-3-229

Moon, R. and Shin, G. (2018). From brain drain to brain circulation and linkage. Stanford, CA: Freeman Spongli Institute of International Studies.

Nair, M., & Webster, P. (2013). Health professionals’ migration in emerging market economies: Patterns, causes and possible solutions. Journal of Public Health (United Kingdom). https://doi.org/10.1093/pubmed/fds087

Ohr, S. O., Jeong, S., Parker, V., & McMillan, M. (2014). Organizational support in the recruitment and transition of overseas-qualified nurses: Lessons learnt from a study tour. Nursing and Health Sciences. https://doi.org/10.1111/nhs.12085

Oladeji, B. D., & Gureje, O. (2016). Brain drain: a challenge to global mental health. BJPsych. International. https://doi.org/10.1192/s2056474000001240

Schumann, M., Maaz, A., & Peters, H. (2019). Doctors on the move: A qualitative study on the driving factors in a group of Egyptian physicians migrating to Germany. Globalization and Health. https://doi.org/10.1186/s12992-018-0434-x

Sheikh, A., Naqvi, S. H. A., Sheikh, K., Naqvi, S. H. S., & Bandukda, M. Y. (2012). Physician migration at its roots: A study on the factors contributing towards a career choice abroad among students at a medical school in Pakistan. Globalization and Health. https://doi.org/10.1186/1744-8603-8-43

Slowther, A., Lewando Hundt, G. A., Purkis, J., & Taylor, R. (2012). Experiences of non-UK-qualified doctors working within the UK regulatory framework: A qualitative study. Journal of the Royal Society of Medicine. https://doi.org/10.1258/jrsm.2011.110256

Subedi, P., Aylott, J., Khan, N., Shrestha, N., Lamsal, D., & Goff, P. (2020). “Hybrid” medical leadership emergency medicine training for international medical graduates. Leadership in Health Services. https://doi.org/10.1108/LHS-05-2020-0027

Takemura, T., Kielmann, K., & Blaauw, D. (2016). Job preferences among clinical officers in public sector facilities in rural Kenya: A discrete choice experiment. Human Resources for Health. https://doi.org/10.1186/s12960-015-0097-0

The Foundation Programme (2019). UK Foundation Programme 2019 F2 career destinations. Available at: https://foundationprogramme.nhs.uk/resources/reports/

The Health Foundation, The King’s Fund and Nuffield Trust (2018). The health care workforce in England: make or break? Available at: https://www.kingsfund. org.uk/publications/health-care-workforce-england

THET (2019). From Competition to Collaboration Ethical leadership in an era of health worker mobility. Available at file:///X:/From-Competition-to-Collaboration_THETPolicyReport-1.pdf

Tiffin, P. A., Illing, J., Kasim, A. S., & McLachlan, J. C. (2014). Annual Review of Competence Progression (ARCP) performance of doctors who passed Professional and Linguistic Assessments Board (PLAB) tests compared with UK medical graduates: National data linkage study. In BMJ (Online). https://doi.org/10.1136/bmj.g2622

Trewby, P. (2017). Migrating doctors. In Clinical Medicine, Journal of the Royal College of Physicians of London. https://doi.org/10.7861/clinmedicine.17-1-4

Webb, J., Marciniak, A., Cabral, C., Brum, R. L., & Rajani, R. (2014). A model for clinical attachments to support international medical graduates. BMJ. https://doi.org/10.1136/bmj.g3936

World Health Organisation. (2016). Health workforce and services Draft global strategy on human resources for health: workforce 2030 Report by the Secretariat.

World Health Organization (2019). Human resources for health - WHO Global Code of Practice on the International Recruitment of Health Personnel: third round of national reporting. Available at: http://apps.who.int/gb/ebwha/pdf_files/WHA72/A72_23-en.pdf.

